# COVID transmission and contact tracing using WHO risk assement tool among frontline healthcare workers : Insights from a South Indian tertiary care centre

**DOI:** 10.1101/2021.04.07.21255044

**Authors:** Merlin Moni, Kiran G Kulirankal, Preetha Prasanna, Ann Mary, Elizabeth Mary Thomas, Rejitha P Sundaram, Binil Babu, Veena Bindu, Fabia Edathadathil, Sai Bala Madathil, K V Beena, Dipu T Sathyapalan

## Abstract

**Background:** The high exposure risk to COVID among frontline heathcare workers was a major challenge to healthcare systems across the globe that warranted close monitoring through risk assessment and contact tracing strategies. The objective of our study was to characterize exposure risk factors for transmission and subsequent COVID positivity among the frontlinehealthcare workers in our institution during the pandemic period.

**Methods:** The retrospective observational study conducted over a period of 6 months from June 2020 to November 2020 at a 1300-bedded South Indian tertiary care centre included frontline healthcare workers who were assessed for their identified encounter with COVID positive individual using a modified WHO COVID risk assessment tool. Additional risk attributes of exposure characterized among COVID positive healthcare workers comprised of shared space, cluster related transmissions and multiple instances of exposure to COVID.

**Results:** Among a total of 4744 contacts with COVID positive individuals assessed for risk stratification during the study period, 942 (19.8%) were high risk and 3802 (80.2%) were low risk exposures respectively. 106 (2.2%) turned COVID positive during the surveillance period of 14 days. Frontline workers working in COVID areas had significant low COVID rates as compared to other areas (N=1, 0.9%). The average monthly COVID positivity rates being 1.66%, the attack rates among high risk and low risk contacts among the total HCWs screened were 5% (46/942) and 1.57% (60/3802) respectively. Shared space (70%) and IPC breaches (66%) were found to be highly prevalent in the COVID positive cohort, along with maskless encounters (43%) and multiple exposure (39%). The attack rate among the 6 identified COVID cluster groups (5.5%) were found to be higher than the attack rate (2.2%) noted among the total contacts screened and no significant association was observed between risk categories in the clusters.

**Discussion:** Our study highlights higher risk of COVID positivity among high risk contacts as compared to low risk contacts. However, the high COVID positivity rate in low risk group among cluster transmissions and its lack of association with risk assessment highlight the suboptimal utility of the risk assessment strategy among cluster groups.

## Introduction

The trajectory of COVID-19 cases have been on the rise ever since the first case in India was detected in Kerala on January 27, 2020^1^. The increased burden of COVID infections among healthcare workers (HCWs) has been evidenced by a prevalence of 2747 infections in 100000 HCWs relative to 242 infections in 100000 in normal community and number of health care workers infected crossed 570,000 with a death toll of more than than 2,500 deaths^2,3^. With the increase in the number of COVID cases, the potential risk for the healthcare workers catering to the state’s high risk, travelling community has also increased. So far, reports show that more than 5000 health care workers have been infected by this virus in this state^4^. (New Indian Express).

The challenges involved in the management of healthcare related COVID were multipronged^5^. Frontline health care workers who are directly involved in the patient care were deduced to be at a higher risk necessitating stringent use of personal protective equipment and work place restrictions including quarantine, staying away from family ranging to frequent surveillance swabbing^6^. Health care institution related outbreaks bore implications like HCW and patient related outbreaks, hospitalisations, mortality and sequelae and even closure of the institution to contain the outbreak^7,8^. Making hospitals safe for patients and HCWS had always been the greatest challenge in this period of time in the background of paucity of data and recommendations in this direction^9,10^. However, relying on quarantine of all contacts of COVID positive individual alone will amount to severe compromise of man hours, which is vital for running hospital services.

The transmission nidus included not only patients but also co-workers posing significant risk of transmission to other health care workers. Sharing of work space and the increased possibility of maskless contact during HCW-HCW interactions owing to the recognized fomite and airborn transmission dynamics of the disease acted as kindlers of spread amongst them^11,6^. In this context, hospitals needed to cater to a heterogenous cross section of population which would includes those who would opt for general Ward and private rooms areas in addition to the common consultation areas. The infrastructural challenge in these areas was the original construction which had aimed to handle high case load in prepandemic times but were not suited to address the challenges of social distancing norms^12^. In addition to this were similar challenges in areas of interactions between the health care workers, as in dining areas and shared space at the out patient areas^11^. All these factors enhanced the exposure risk of HCW at workplace.

With rising rates of community transmission it is imperative that increasing encounters with COVID positive patients from ambulatory and inpatient areas are not uncommon. Ensuring the adherence to the PPE policy, especially mandating N95 masks and other PPE appropriate to the locations based on the risk of the patient population catered to had been the cornerstone of preventing health care associated COVID transmission^13,14^. Barring the hurdles of training of HCWs for the proper use of PPE and provision of PPE, this was supposed to substantially bring down risk of HCW contracting the disease from infected individual. A strategy of categorizing the contacts of COVID positives into ‘High Risk’ Contacts and ‘Low risk Contacts’ based on known exposure factors was formulated by CDC and the same were adopted into formal policy of governments and institutions for the purpose of rational quarantine of the HCWs^15,16^. Despite majority of the institutions following up the risk categorization based on the guidelines, robust data on COVID positivity rates among High and Low risk contacts are sparse to support informed administrative decisions for health policy makers regarding optimal resource utilization of healthcare workers^17^. Moreover, the need of exploring unaccounted exposure variables like clustering in the risk profiling strategy that could impact positivity rates among Low risk contacts is imperative in a healthcare setting catering to COVID patients and potentially opens up the possibility of incorporating hitherto unaccounted exposure factors into the existing risk profiling strategy. The exposure related risk profiling among frontline healthcare workers needs to be further refined from the broader risk profiling methodology pertaining to healthcare workers. We hereby describe our institutional experience in charecterising the risk profile of COVID positive frontline healthcare workers across our 1300-bedded tertiary care academic hospital in South India.

## Methods

### PROCESS OF RISK ASSESSMENT

The risk assessment form was initially designed by incorporating key elements of world Health Organisation(WHO), COVID contact questionnaire, which comprised of basic demographics, co-morbidities, type of PPE worn by the contact and COVID positive individual, exposure time during their encounter, perceived distance maintained during exposure, details of direct contact with resources used for COVID positive patient and Infection prevention & control (IPC) breaches observed^18,15^. However the questionnaire were tailored to our setting by adding questions to assess social interactions and distinct behavioural practices.

Two distinct questionnaires were incorporated for contact with COVID positive patient and COVID positive HCW respectively to capture the IC practices and breach in practicing. The questions assessed the occurrence of conversations, close contact or shared space with the COVID positive individual. The encounters were assessed by interviews over the phone. The risk assessment forms are included in Annexure 1.The Command centre comprising of administrative champions, Infectious Disease physicians, infection control nurses and other departments formed the multidisciplinary committee in charge of the process. The risk assessment forms were made available in shared folder in all desktops of the hospital for easy access. The duly filled forms were required to be submitted to the Infection Control department. A panel of doctors with training and experience in infection control evaluated the submitted forms and the respondents were divided into high and low risk for COVID. HCW belonging to low risk category were advised self-monitoring for symptoms and allowed to resume work. High risk category was advised a quarantine period of 14 days file according to State government revised guidelines for COVID testing and quarantine. Retrospective review of risk assessment forms submitted to infection control department at the time of identified contact with COVID positive individual. The medical panel evaluated the components of risk stratification in all cases and categorized into high risk exposure and low risk exposure. For the risk stratification the prime factor considered was the appropriateness of PPE in the given setting as per the institutional protocol.

### Data dictionary

#### Health personnel definitions

##### Healthcare worker

all paid and unpaid persons serving in healthcare settings who have the potential for direct or indirect exposure to patients or their infectious aerosols, secretions and materials (e.g., doctors, nurses, laboratory workers, facility or maintenance workers, clinical trainees, volunteers).

##### Frontline health care workers

Healthcare worker contacts who are directly exposed to the patient as part of their routine clinical work such as doctors, nurses or health-care workers as participants who reported direct patient contact.

#### Risk factor definitions

##### Shared space

A close proximity of the HCW contact with the COVID-19 confirmed patient attributable to social interactions such as shared dining, living or work spaces and routine meetings for clinical case discussions or handovers.

##### Maskless encounter

HCW contact encounters with COVID-19 confirmed patient or other HCW in which either the HCW or the COVID-19 confirmed individual was not wearing appropriate facemask recommended.

##### Cluster transmissions

A cluster of cases was defined as 2 or more cases of SARS-CoV-2–positive COVID-19 among HCWs who work in the same unit area at overlapping times^19^.

##### Time of contact

Face to face contact with a probable or confirmed case for at least 15 minutes^20,21^

##### IPC breach

Perceived IPC breaches by HCWs with respect to appropriate PPE use or standard IPC practices.

##### Average distance during encounter

Direct contact with with a probable or confirmed case who may have been within 1 metre of the healthcare worker.

## Results

Over a period of 6 months from June 2020 to November 2020, 4744 instances of recorded contact with COVID positive individuals were assessed for risk stratification as per the WHO guidelines 942 (19.8%) high risk encounters were identified among the 4744 instances and 3802 (80.2%) were categorized as low risk.

### Baseline characteristics

Among the 4744 contacts traced as part of the risk assessment process, 106 front line health workers(2.2%) turned COVID positive (Table 1) including doctors, nurses and nursing assistants. The positivity was more in females (76%,n=81). Nurses were the major occupational category class with maximum positivity among the contacts (48%, n=51).

**Table 1:**
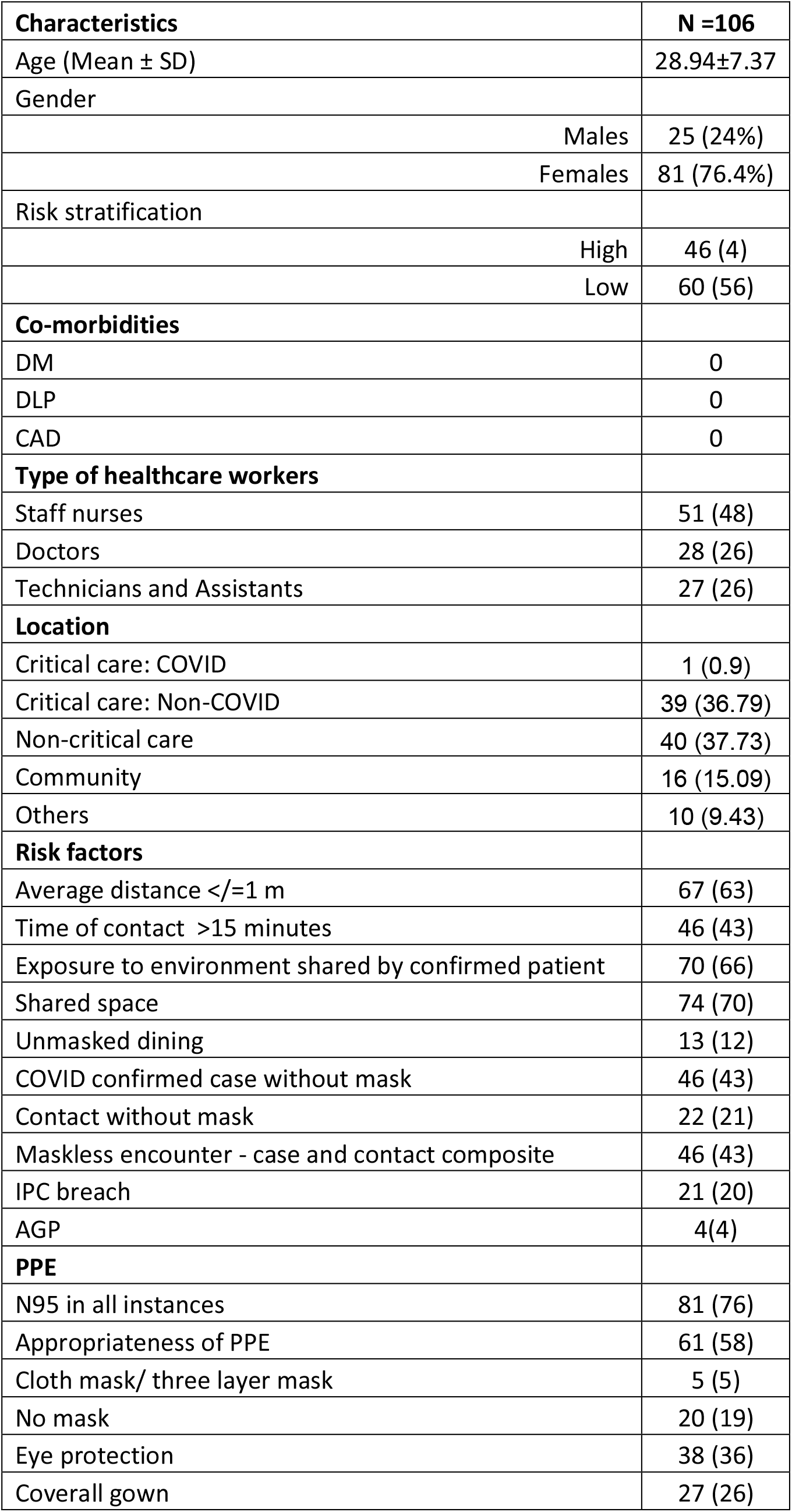
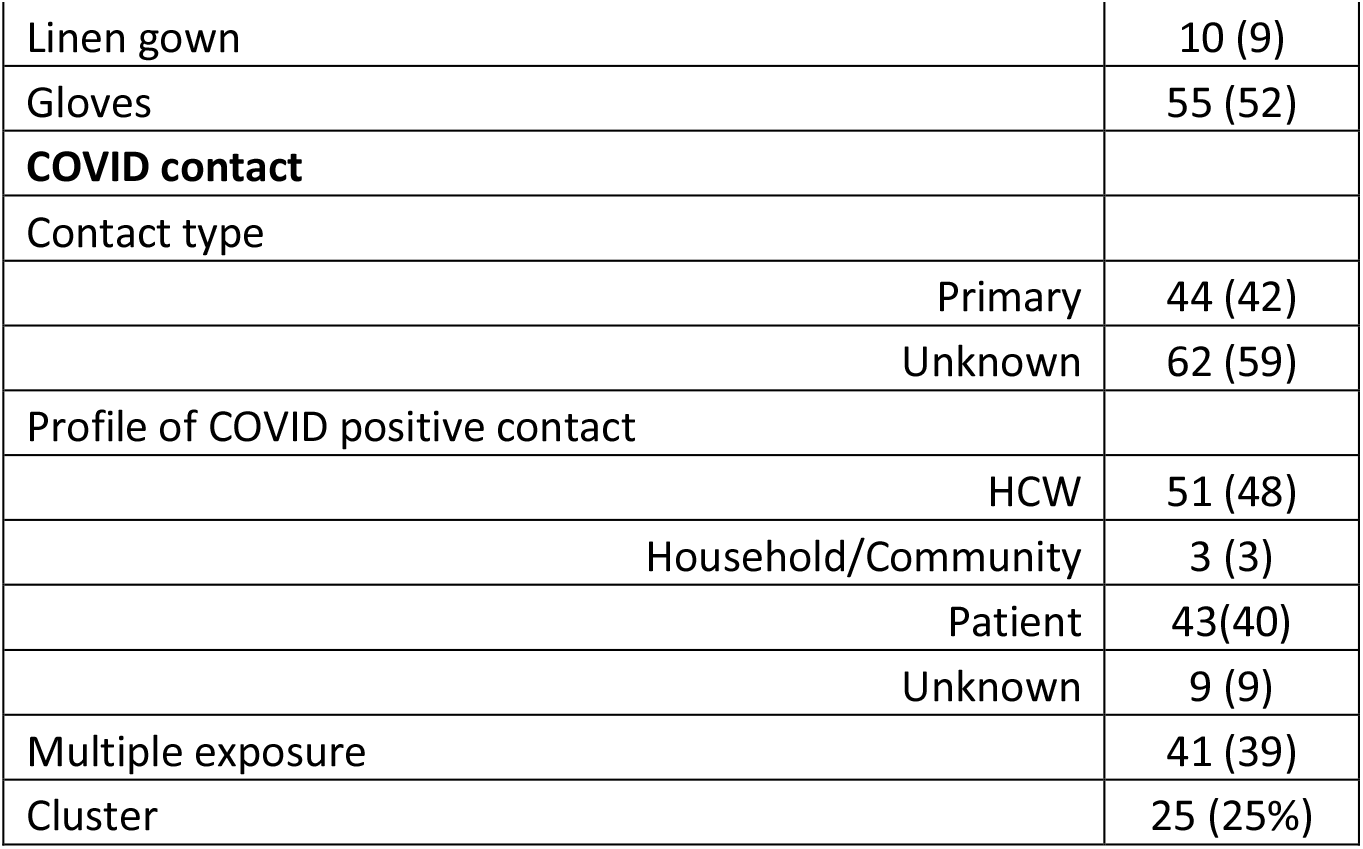
Baseline characteristics of COVID positive patients.

| Characteristics | N =106 |
| --- | --- |
| Age (Mean ± SD) | 28.94±7.37 |
| Gender |  |
| Males | 25 (24%) |
| Females | 81 (76.4%) |
| Risk stratification |  |
| High | 46 (4) |
| Low | 60 (56) |
| **Co-morbidities** |  |
| DM | 0 |
| DLP | 0 |
| CAD | 0 |
| **Type of healthcare workers** |  |
| Staff nurses | 51 (48) |
| Doctors | 28 (26) |
| Technicians and Assistants | 27 (26) |
| **Location** |  |
| Critical care: COVID | 1 (0.9) |
| Critical care: Non-COVID | 39 (36.79) |
| Non-critical care | 40 (37.73) |
| Community | 16 (15.09) |
| Others | 10 (9.43) |
| **Risk factors** |  |
| Average distance ≤/=1 m | 67 (63) |
| Time of contact >15 minutes | 46 (43) |
| Exposure to environment shared by confirmed patient | 70 (66) |
| Shared space | 74 (70) |
| Unmasked dining | 13 (12) |
| COVID confirmed case without mask | 46 (43) |
| Contact without mask | 22 (21) |
| Maskless encounter - case and contact composite | 46 (43) |
| IPC breach | 21 (20) |
| AGP | 4(4) |
| **PPE** |  |
| N95 in all instances | 81 (76) |
| Appropriateness of PPE | 61 (58) |
| Cloth mask/ three layer mask | 5 (5) |
| No mask | 20 (19) |
| Eye protection | 38 (36) |
| Coverall gown | 27 (26) |
| Linen gown | 10 (9) |
| Gloves | 55 (52) |
| **COVID contact** |  |
| Contact type |  |
| Primary | 44 (42) |
| Unknown | 62 (59) |
| Profile of COVID positive contact |  |
| HCW | 51 (48) |
| Household/Community | 3 (3) |
| Patient | 43(40) |
| Unknown | 9 (9) |
| Multiple exposure | 41 (39) |
| Cluster | 25 (25%) |

Major stake of exposures resulting in positivity were traced to other health care workers (n=51, 48% followed by COVID positive patients (n=43, 40%). More than half of the patients had single COVID traceable contact identified in the prior 2 weeks of COVID positivity (n=53, 50%). Almost equal number of patients had multiple identified contacts n=41, 32% or non-traceable contacts(n=9, 9%). Only a single COVID positive case (n=1, 0.9%) has been reported from the COVID critical care area.

Among 942 high risk contacts, 46 (4.88%) turned out to be positive and among the 3802 low risk contacts, 60 turned COVID positive (1.5%) on prospective surveillance. The attack rate among high risk and low risk contacts among the total HCWs screened during the study period were 5% (46/942) and 1.57% (60/3802) respectively.

### Description of IPC practices

22 among 106 HCW (21 %) were traced to have maskless encounter with the COVID positive individuals. In 46 instances, the COVID positive individual were not wearing mask during encounter. In 20 instances neither the HCW nor the COVID positive individual was wearing a mask. In the cohort of 106 HCWs, 81 (76%) wore N95 masks during encounter with COVID positive individual. Eye protection in the form of goggles or visors were worn in 38 instances (36%). Gloves for contact precautions were worn in 55 instances (52%). Overall the appropriateness of the used PPE was estimated to be 57.5% (n=61). A self-perceived breach in IPC practice was identified in 21 instances (20%).

### Exposure details

Only 4 out of 106 (3.7%) affected frontline workers reported doing Aerosol generating procedures for COVID positive patients. Non healthcare related activities like unmasked dining among health care workers were identified in 13 out of 37, (35%) instances of contracting COVID from other health care workers. Time of contact more than 15 minutes was observed in 46 instances (n= 46, 43%). Encounter with identified COVID positive individual at a distance less than 1 meter was reported in 63% (n=67). The distribution of identified risk attributes including multiple exposures, cluster and shared space among COVID positive cohort are depicted in Fig 1A and Fig 1B.

**Fig 1A:**
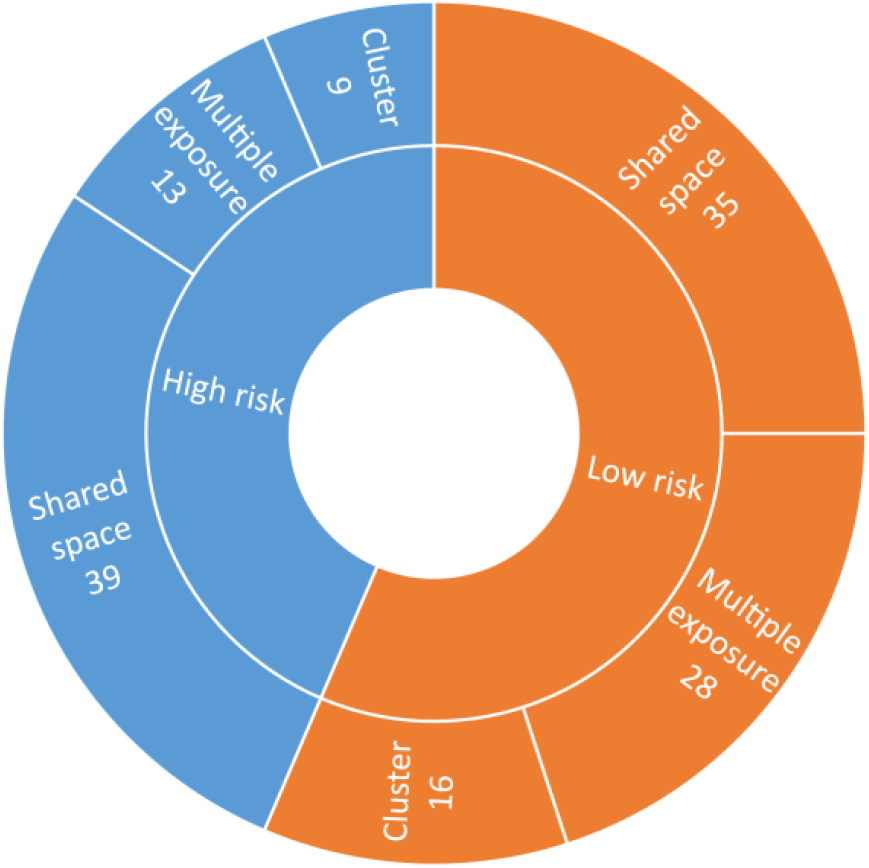
Distribution of major transmission risk factors among WHO risk categories.

**Fig 1B:**
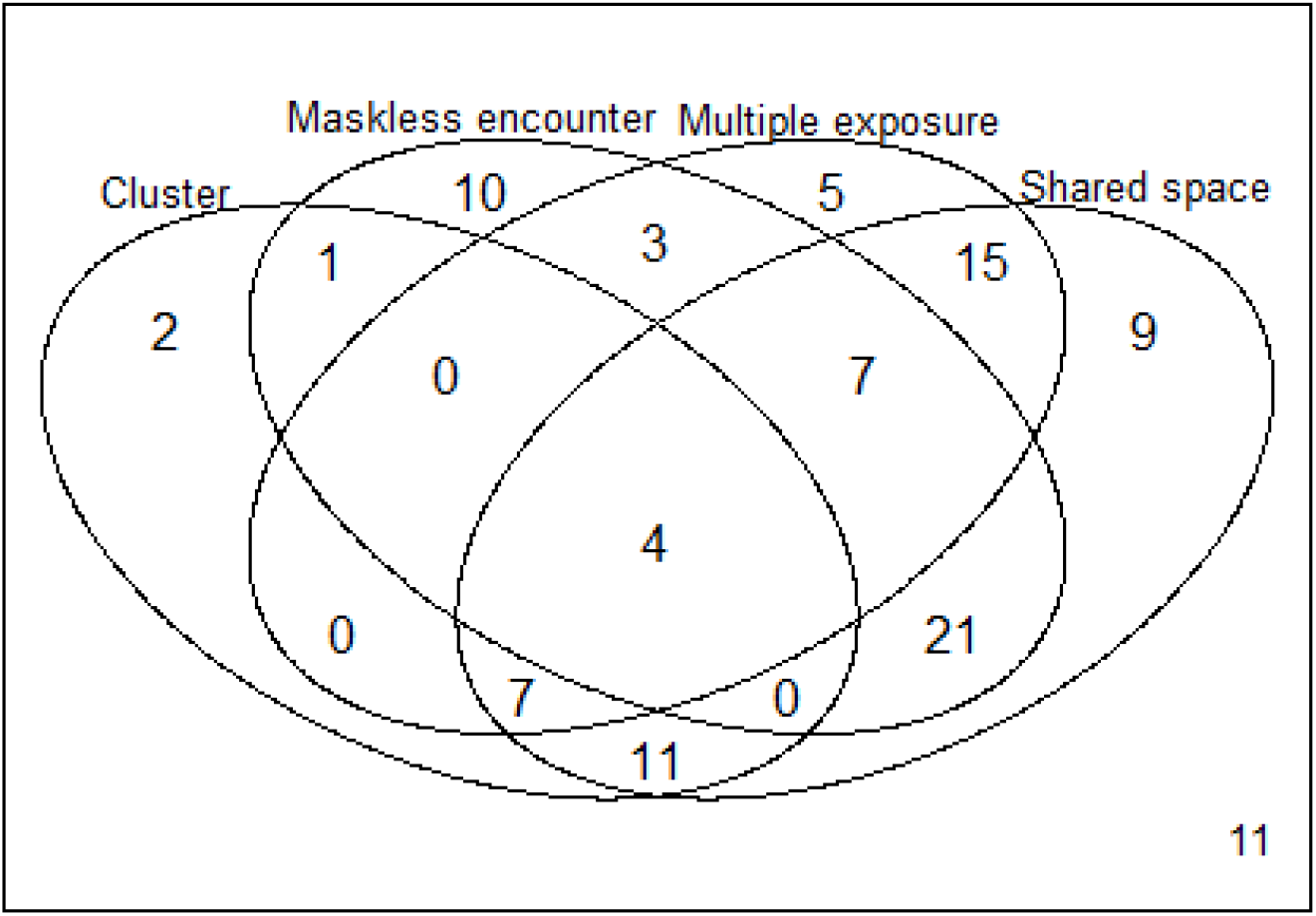
Venn diagram representing the distribution of risk factors among COVID positive group.

### COVID positivity rates

Fig 2 depicts the monthly distribution of COVID positivity rates from June to November 2020 among HCWs screened as part of risk assessment. Average monthly COVID positivity rate during the study period was 1.66%. Among high risk and low risk HCWs, the average monthly COVID positivity rate were observed to be 5.76 and 1.05 respectively. Among high risk HCW group, the monthly COVID positivity rate exhibited a sharp rise from 2.02% in August 2020 to 20.5% in the month of November 2020. The COVID positivity rate among low risk HCW group demonstrated a gradual increase from 0.87% in August to 2.73% in November 2020.

**Fig 2:**
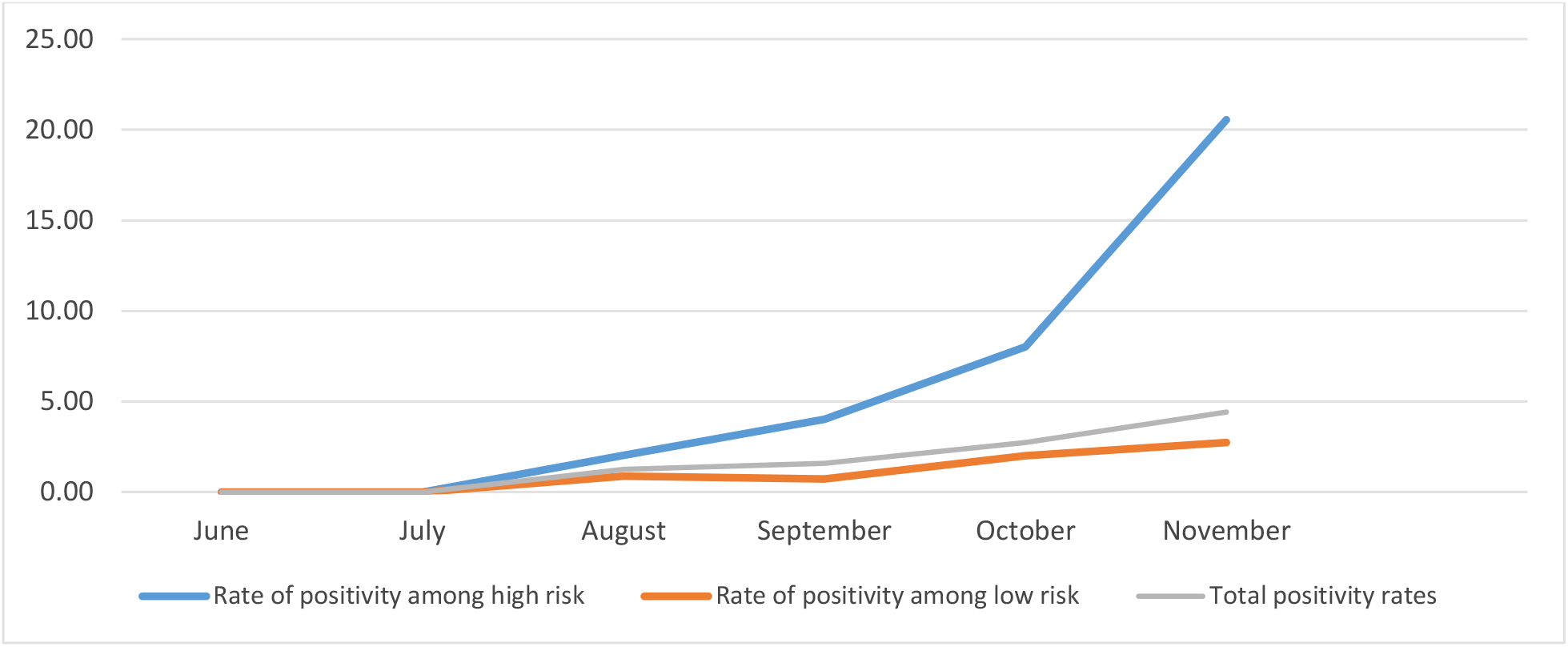
Distribution of monthly COVID positive rates.

### Cluster transmissions

During the study period, 6 groups of cluster transmission were identified in our healthcare setting comprising of 25 COVID positive HCWs among 489 screened contacts. The largest number of confirmed cases in a single cluster was 8. Among the low risk category, COVID positive cases in cluster transmissions accounted for 27% (n=16).

The distribution of COVID positive cases among high risk and low risk groups in cluster transmissions are depicted in Fig 3. The total attack rate among cluster groups was estimated to be 5.11% among cluster transmissions. The attack rates were observed to be 6.87% and 4.53% in high risk and low risk groups respectively. A significant association was not observed between COVID positivity and risk assessment categories among cluster groups (p = 0.3).

**Fig 3A:**
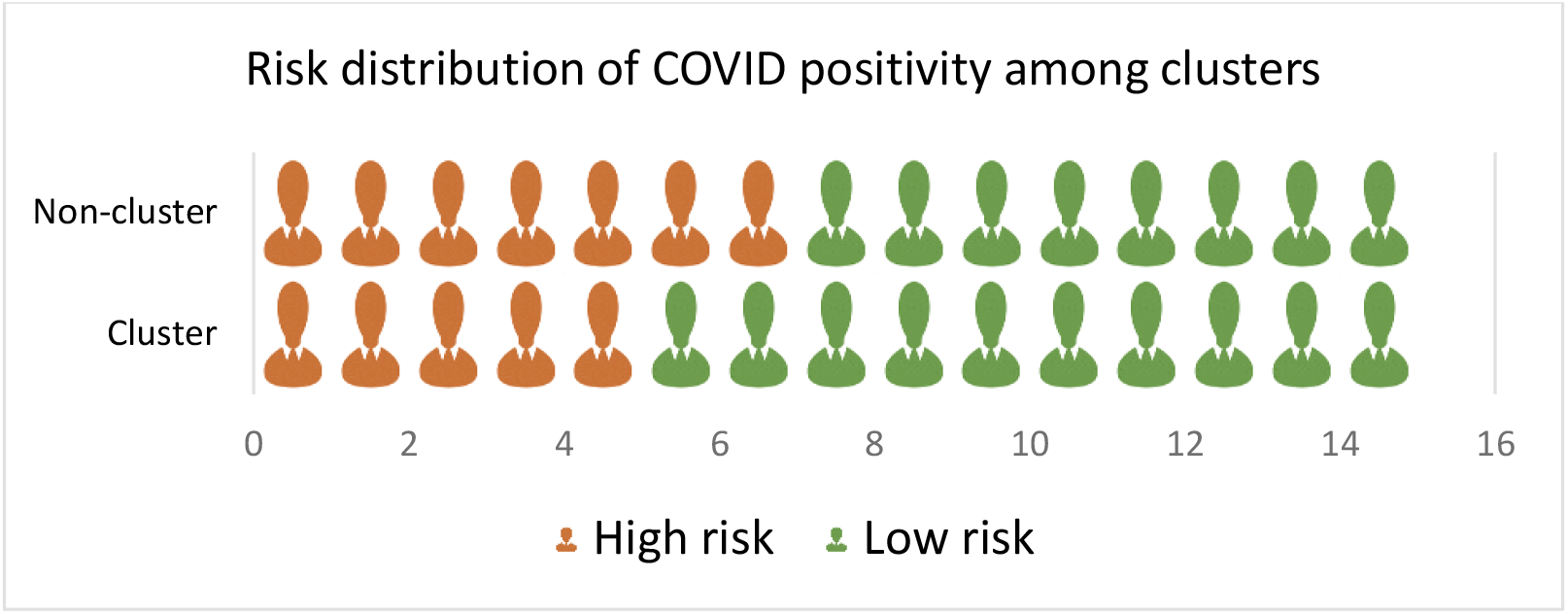
Infographics depicting the distribution of clusters among COVID positive cohort.

**Fig 3B:**
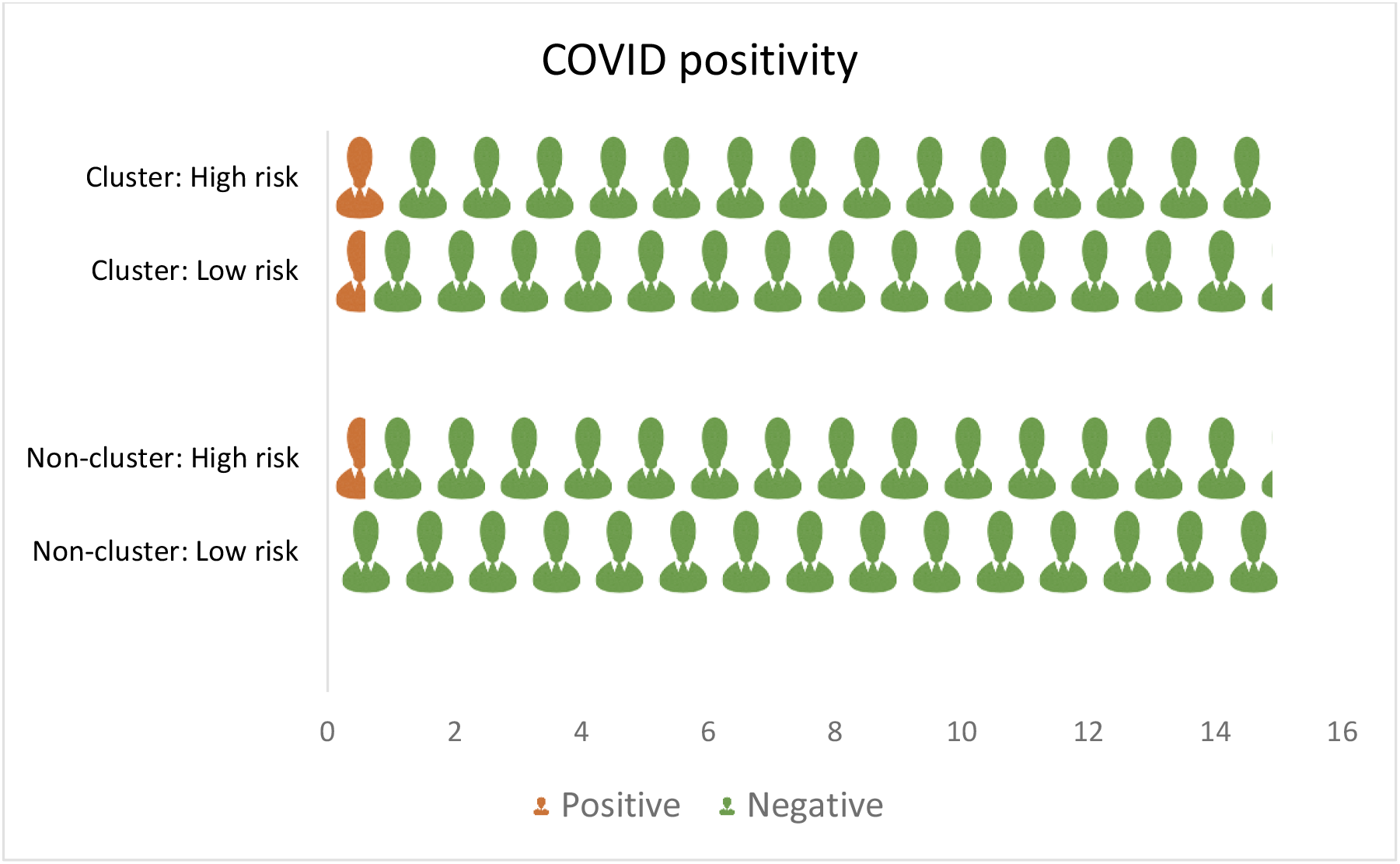
Infographics depicting the distribution of clusters among the whole cohort.

## Discussion

Our research study characterized the exposure risk and subsequent COVID positivity rate among the front line workers of our instituition on following the WHO COVID contact questionnaire tailored to our setting. WHO COVID contact risk assesement questionnaire is followed world wide for healthcare worker exposure risk assessment and quarantine which helped to decrease the quarantine of health care workers and saved man hours at work. However the application of the tool in predicting the attack rates among the frontline health care workers was less widely explored.

The average attack rate among high risk contacts (5%) was found to be more than three times(1.5%) that of the low risk contact, with peak attack rates of upto 20% and 2.5% among high risk and low risk contacts repsectively.This amounts to an attack rates ranging from from 1:3 to 1:8 among low risk contacts as compared to high risk contact. In our study, COVID positivity rates have been comparable among clinicians and nurses (4%) as compared to higher COVID positivity rates among nurses (21%) in comparison to doctors(13%) in a recent Indian study^22,23^ which could be probably due to the better adherence to the infection control practices in our instituition.

COVID positivity was observed to be negligible in COVID critical care areas as compared to non COVID areas where PPE policies had been strictly adhered to, underlining the importance of an effective PPE policy in thwarting the exposure risk. The HCW positivity was more likely to be acquired from other HCWs sharing the same space rather than a patient related exposure who has shared the same space^22^. The findings from our study highlights the need for policies to avoid maskless activities like dining together and to enhance the protection in shared spaces with usage of mask and social distancing.

Our research study earmarked major exposure risk variables comprising of shared space, IPC breaches and maskless encounters in addition to characterization of cluster transmissions and multiple exposure profiles of COVID positive cohort among our frontline healthcare workers. Considering the negligible rates of COVID acquisition among healthcare workers from COVID critical care areas of our instititon as demonstrated by our study, avoiding of unnecessary quarantine of HCWs working in COVID positive areas could be pursued, owing to the the low risk of contracting COVID from such areas than those working in non COVID areas, in the setting of high community prevalence. Infrastructural changes involving the redesigning of shared spaces within the healthcare setting has to be considered, owing to the greater stake of “shared space >15 mts” in HCW covid positivity and cluster transmissions. Staff dining areas are to be refurbished to avoid face to face maskless encounters and to maintain social distancing.

Monthly distribution of COVID positivity rates among high risk HCW group approximately doubled per month from August to November, primarily owing to the surge in community transmissions during the peak outbreak phase of COVID pandemic in 2020. This depicts the interplay of multiple factors especially the pivotal influence of community transmission rates on inhospital HCW transmission risks.

A major proportion of COVID positive HCWs who were part of the cluster transmissions were categorized as low risk during the initial risk assessment process, accounting for 64% (16/25) of COVID positive HCWs in the cluster groups. Cluster tranmsissions are increasingly characterized by shared environments, possibility of multiple unidentified exposures, high incidence of social interactions involving individuals belonging to multiple spheres of care comprising of care givers, HCWs and patients themselves. The actual probability of being positive in a classical low risk contact as measured by the exposure risk among an identified cluster can thus cross over to that of a high risk contact. The higher probability among cluster transmissions for the incidence of COVID positive cases was unidentified during the routine risk assessment process.The attack rate among COVID cluster groups (5.5%) were observed to be higher than the attack rate (2.2%) noted among the total contacts screened as part of the risk assessment. Similarly, attack rates were also found to be higher among the high(6.8%) and low risk groups (4.5%) in the cluster transmissions relative to the overall high(5%) and low risk groups (1.57%) of the total contacts screened. Among the total contacts screened, the COVID positiviy rates of high risk group was significantly higher than that of low risk group (p<0.001). Despite a slightly higher COVID positivity of 6.87% in high risk group in comparison to 4.35% in low risk group in cluster based infections, no significant association was observed between the COVID positivity and risk assessment groups among cluster transmissions. This would potentially highlight the suboptimal utility of the risk assessment strategy among cluster transmissions. This also calls in for location based surveillance in healthcare settings for early detection of cluster formation and tailored contact tracing in case of cluster transmissions owing to the higher attack rates.

### LIMITATIONS

The causality of the identified co-variates could not be determined as data regarding COVID negative contacts were not available.

## Data Availability

The data is available with the corresponding author upon reasonable request

